# Serum brain-derived p-tau 217 and SV2A Reduce Peripheral Interference in Alzheimer’s Disease: A Multicohort Study

**DOI:** 10.64898/2026.07.28.26359122

**Authors:** Mingjie Ma, Hongxu Wang, Xinghao Lian, Jinyue Li, Ninglu Gao, Jie Liu, Xiaohan Sun, Kunli Wang, Jingwen Xu, Kai Shao, Bing Zhao, Fang Yuan, Cuiping Zhao, Ou Chen, Wei Li, Yuying Zhao, Yi Li, Chuanzhu Yan, Shuangwu Liu, Fuchen Liu

**Affiliations:** Department of Neurology, Shandong Key Laboratory of Mitochondrial Medicine and Rare Diseases, Research Institute of Neuromuscular and Neurodegenerative Disease, Qilu Hospital of Shandong University, Jinan, Shandong, China; Department of Neurology, Qilu Hospital (Qingdao), Cheeloo College of Medicine, Shandong University, Qingdao, China; Department of Neurology, Zhongshan Hospital (Xiamen), Fudan University, Xiamen, China; Department of Neurology, The Fourth Hospital of Jinan City, Jinan, China; Department of Medicine Experimental Center, Qilu Hospital (Qingdao), Cheeloo College of Medicine, Shandong University, Qingdao, Shandong, China; School of Nursing and Rehabilitation, Cheeloo College of Medicine, Shandong University, Jinan, China

**Keywords:** AD, ALS, p-tau 217, brain-derived p-tau 217, SV2A

## Abstract

**Importance:** Recently, increasing studies have demonstrated that blood p-tau 217, the most promising diagnostic biomarker for Alzheimer’s disease (AD), is increased and originated from muscle damage in amyotrophic lateral sclerosis (ALS) patients. These findings suggested that blood total p-tau 217 may partly derive from muscle damage. Thus, there is an urgent need for identifying blood brain-derived p-tau 217, but not blood total p-tau 217 which may contain muscle-derived p-tau 217, and other brain-derived biomarkers in order to reduce the potential peripheral interference in its adoption in assisting early diagnosis in AD patients.

**Objective:** To explore whether serum brain-derived p-tau 217 is a more promising diagnostic biomarker and has less peripheral interference than total p-tau 217 for AD patients in a large multicentre cohort. To examine whether serum synaptic vesicle glycoprotein 2A (SV2A) is a potential biomarker for assessing brain damage in AD patients. To examine whether serum p-tau 217 is a specific lower motor neuron (LMN) damage biomarker for ALS patients.

**Design, Setting, and Participants:** This cross-sectional study was conducted in 3 independent cohorts and a total of 1198 participants, including 325 AD patients, 235 ALS patients, 289 LMN disease controls (LMNDCs), 145 dementia controls (DDCs), and 204 cognitive intact healthy controls (CIHCs).

**Main Outcomes and Measures:** Serum brain-derived p-tau 217, total p-tau 217, SV2A, and NfL were measured based on single molecular detection technique.

**Results:** Serum brain-derived p-tau 217 was significantly increased in AD patients compared to ALS patients, DDCs, LMNDCs and CIHCs, while serum p-tau 217 was significantly increased in both ALS patients and AD patients compared to DDCs, LMNDCs and CIHCs after familywise error correction (*p* < 0.05). Serum SV2A was significantly decreased in AD patients than in other groups. Moreover, area under the curve for serum brain-derived p-tau 217 in differentiating AD from other groups were 0.927-0.954.

**Conclusions and Relevance:** Our findings suggest that serum brain-derived p-tau 217 and SV2A are more specific diagnostic biomarkers for reflecting brain damage and may not be disturbed by peripheral damage for AD patients. Moreover, we suggest that serum p-tau 217 is a specific LMN damage biomarker for ALS patients.

**Key Points:** *Question:* Whether blood brain-derived p-tau 217 is a more promising diagnostic biomarker and has less peripheral interference for Alzheimer’s disease (AD) patients than blood total p-tau 217, which may partly originate from muscle damage.

*Findings:* Serum brain-derived p-tau 217 was significantly increased in AD patients compared to amyotrophic lateral sclerosis (ALS) patients, other lower motor neuron disease controls (LMNDCs), dementia controls (DDCs), and cognitively intact healthy controls (CIHCs), while serum p-tau 217 was significantly increased in both ALS patients and AD patients compared to DDCs, LMNDCs and CIHCs.

*Meaning:* Our findings first suggest that blood brain-derived p-tau 217 is likely a more ideal diagnostic biomarker with less peripheral interference than total p-tau 217 for AD. **Statistical analysis:** Statistical analyses were performed by Dr. Xiaohan Sun

## Introduction

With the increasing severity of population aging, Alzheimer’s disease (AD) has become a huge public health issue, imposing a heavy burden on society and the families of patients.^1–3^ Specifically, early diagnosis is conducive to the early treatment of AD and have a crucial role in delaying the time window for patients to enter the stage of cognitive deficits and dementia.^2–11^

At present, according to the newly AD diagnostic criteria and published studies, blood phosphorylated tau 217 (p-tau 217) is core 1 and the one of the most promising blood biomarkers for assisting early diagnosis in AD patients, which may reflect brain amyloid damage.^1–11^ However, recently, several studies have found that blood p-tau 217 are increased and derive from muscle damage, but not related to AD pathology, in amyotrophic lateral sclerosis (ALS) patients, which is largely different from AD patients.^12–15^

Importantly, although previous studies have demonstrated that elevated blood p-tau 217 are peripheral sources and unrelated to β-amyloid (Aβ) pathology in ALS, and AD patients and ALS patients show significant differences in clinical symptoms, these findings remain implied that blood total p-tau 217 may partly derive from muscle damage in some participants and still pose some challenges for using p-tau 217 in assisting early diagnosis in AD patients, in particular in those elderly in community.^6–15^

Thus, there is an urgent need for identifying blood brain-derived p-tau 217, but not blood total p-tau 217 which may contains muscle-derived p-tau 217, in order to reduce the potential peripheral interference in its adoption in assisting early diagnosis in AD patients.^7–16^ Importantly, a novel brain-derived biomarker, that is brain-derived p-tau 217, an antibody engineered to specifically target tau isoform originating from the brain as detection antibody and paired with p-tau217 antibody as capture antibody to form a sandwich assay.^16–18^ To date, several studies have demonstrated that brain-derived p-tau 217 has better performance in assisting early diagnosis in AD patients.^16–18^

However, to our knowledge, no large multicentre study has examined the diagnostic performance of blood brain-derived p-tau 217 in AD patients to date.^16–18^ Moreover, although blood brain-derived p-tau 217 is likely to specifically reflect brain damage, but not peripheral damage in health and disease conditions, no previous study has examined whether serum brain-derived p-tau 217 is elevated or not in patients with ALS and other peripheral damage diseases to date.^11–18^

Specifically, based on the ATN framework, biomarkers reflecting neurodegeneration may also be important for assisting early diagnosis and reducing the potential peripheral interference in AD patients.^3, 19^ Specifically, synaptic vesicle glycoprotein 2A (SV2A) is selectively localized to the membranes of presynaptic vesicles at neuronal axon terminals and currently represents as the reliable *in vivo* biomarker capable of quantitatively assessing cortical synaptic density with high spatial specificity and precision.^20, 21^ Recently, increasing studies have demonstrated that blood SV2A is likely a promising biomarker to uncovers cortical synapse loss and reflecting brain damage in AD patients.^20^ However, to date, no large multicentre study has examined blood SV2A alterations and its diagnostic performance in AD patients.^20^ Moreover, to date, only one large multicentre study has explored blood total p-tau 217 alteration in ALS patients, and that study did not include other lower motor neuron (LMN) disease patients.^12–15^

Moreover, to our knowledge, very few previous studies have examined blood p-tau 217 alterations in other lower motor neuron disease patients to date.^12–15^ Thus, whether blood p-tau 217 can be detected in other lower motor neuron disease patients, and whether blood p-tau 217 is a specific biomarker for assessing LMN damage in ALS patients remain largely unclear.^12–15, 22^

Thus, in the current study, we included three cohorts and a total of 1198 participants, including 325 AD patients, 235 ALS patients, 289 other lower motor neuron disease controls (LMNDCs), 145 dementia controls (DDCs), and 204 cognitive intact HCs (CIHCs), and serum total p-tau 217, brain-derived p-tau 217, NfL, and SV2A levels were measured in all participants based on single molecular detection technique.^9, 20, 23–25^

Specifically, in our large multicentre cohort, we have three aims. First, we aimed to explore whether blood brain-derived p-tau 217 is a potential better biomarker and has less peripheral interference than blood total p-tau 217 for AD patients in distinguishing DDCs, ALS patients, LMNDCs, and CIHCs. Second, we have examined whether blood SV2A is a potential biomarker for assessing brain damage in patients with AD. Finally, we also aimed to examine whether blood p-tau 217 is a specific LMN biomarker and may originate from muscle damage in ALS patients.

## Methods

### Participants

In this study, the inclusion criteria for ALS patients were as follows: 1) met the Awaji criteria for probable or definite ALS.^26^ 2) underwent clinical assessment, serum biomarker measurement, and genetic testing. The exclusion criteria for ALS patients were as follows: 1) refusal to participate; 2) patients who needed nutritional or respiratory support were also excluded; 3) genetic ALS patients based on the genetic testing; 4) comorbidity of other disorders.^27, 28^

For AD patients, patients who met the National Institute on Aging and Alzheimer’s Association criteria (2011) were included, and the diagnosis of AD was supported by the AV-45 PET or CSF Aβ levels in this study.^3, 29^ Patients with Alzheimer’s disease (AD) will be excluded from the study if they refuse to participate, if their AV-45 PET or cerebrospinal fluid (CSF) biomarker test results are Aβ-negative, or if they have other neurological or systemic diseases that may affect cognitive function or biomarker concentrations.^3, 11, 16–18^ In this study, in line with previous studies, other type patients with cognitive deficits with Aβ-were divided into DDCs group.^3–11, 16–18^

Moreover, similar to previous studies, LMNDCs were defined as patients who had chronic progressive weakness or bulbar palsy, such as myasthenia gravis, Kennedy disease, myositis, Duchenne muscular dystrophy (DMD), cervical spondylopathy, and were subjected to the same exclusion criteria as the ALS patients.^28, 30–32^

In the current study, a group of CIHCs, defined as MMSE score was ≥28 and did not report subjective cognitive decline in the current study, were recruited from the community.

Finally, three cohorts of 325 AD patients, 235 ALS patients, 289 LMNDCs, 145 DDCs, and 204 CIHCs were included (Figure 1). In this study, in cohort 1 (Jinan cohort), 163 AD patients, 118 ALS patients, 145 LMNDCs, 71 DDCs, and 102 CIHCs was included from Qilu Hospital of Shandong University, Jinan, Shandong, China. In cohort 2 (Qingdao cohort), 97 AD patients, 70 ALS patients, 86 LMNDCs, 44 DDCs, and 61 CIHCs was included from Qilu Hospital (Qingdao), Shandong University, Qingdao, China. In cohort 3 (Xiamen cohort), 65 AD patients, 47 ALS patients, 58 LMNDCs, 30 DDCs, and 41 CIHCs was included from Xiamen Branch, Zhongshan Hospital, Fudan University, Xiamen, China.

**Figure 1.**
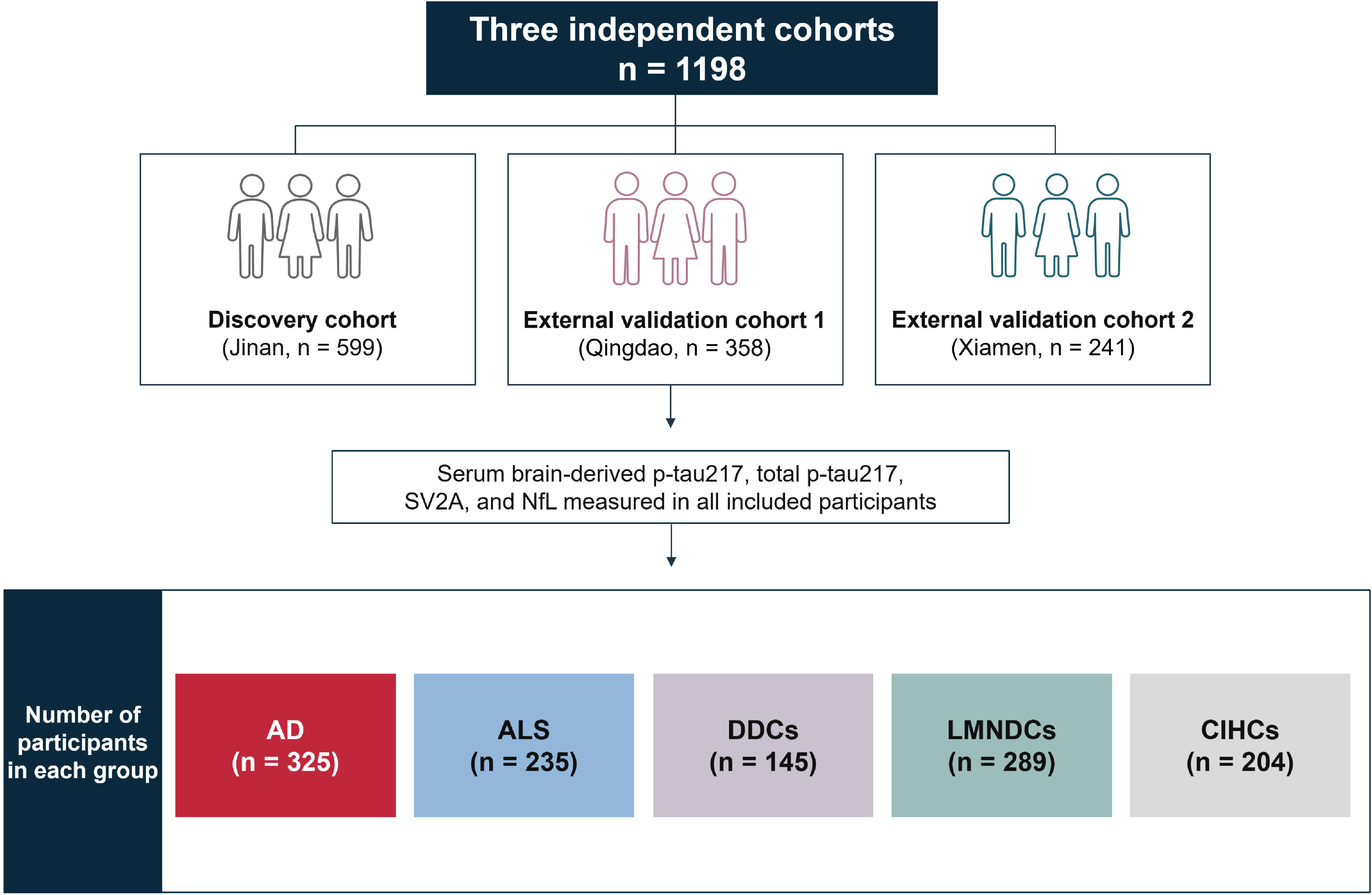
Study Design and Participant Distribution Across 3 Independent Cohorts. A total of 1198 participants were included in the discovery cohort (Jinan; n = 599), external validation cohort 1 (Qingdao; n = 358), and external validation cohort 2 (Xiamen; n = 241). Serum brain-derived p-tau217, total p-tau217, SV2A, and NfL were measured in all participants. The study population comprised 325 participants with AD, 235 participants with ALS, 289 LMNDCs, 145 DDCs, and 204 CIHCs. Abbreviations: AD, Alzheimer disease; ALS, amyotrophic lateral sclerosis; CIHC, cognitively intact healthy control; DDC, dementia control; LMNDC, lower motor neuron disease control; NfL, neurofilament light chain; p-tau217, phosphorylated tau at threonine 217; SV2A, synaptic vesicle glycoprotein 2A.

### Clinical assessments

In this study, ALS patients and LMNDCs’ demographic and clinical information was recorded, including age, sex, education, family history, comorbid conditions, site of symptom onset, and baseline disease duration (time from disease onset to diagnosis).^27, 28^ For ALS patients, disease severity was assessed with the Amyotrophic Lateral Sclerosis Functional Rating Scale-Revised (ALSFRS-R).^28^ Cognitive performances were assessed using Mini-Mental State Examination (MMSE) and Edinburgh Cognitive and Behavioural ALS Screen (ECAS) in all ALS patients, LMNDCs and CIHCs.^28^

Moreover, AD patients’ demographic and clinical information was recorded, including age, sex, education, and disease duration.^21^ Then, MMSE and MoCA were assessed in all AD patients.

### Samples collection and measurements

In this study, in line with our previous studies, all participants completed serum sample collection, and the detailed processes of samples collection and processing have been provided in our previous studies.^28, 33^ All clinical measures were performed within 12 hours of serum sample collection.

In the present study, similar to previous studies, serum brain-derived p-tau 217, p-tau 217, SV2A and NfL levels were measured based on single molecular immunity detection technique in all participants.^9, 20, 23–25^

Moreover, serum p-tau 217 levels and NfL levels were also measured based on Simoa approach in a subgroup of 461 participants.

## Ethics approval

The study was approved by the institutional review boards of Qilu Hospital of Shandong University (KYLL-202412-067-1), Qilu Hospital (Qingdao) of Shandong University (KYLL-202310-016-2), and Zhongshan Hospital (Xiamen), Fudan University (B2024-059R). All participants provided written informed consent.

## Statistical analysis

Continuous variables are reported as the means and standard deviations, and categorical variables are reported as frequencies and proportions. Student’s *t*-tests or analysis of variance (ANOVA) were used to compare continuous variables (with Mann–Whitney *U* tests if necessary). Categorical variables were compared using chi-squared tests. Familywise error (FWE) correction was used, and values of *p* < 0.05 indicated significance. Moreover, diagnostic performances of the serum biomarkers were assessed using the receiver operating characteristic (ROC) curve. The AUC was applied to summarize the classification accuracy of diagnostic models, and 95% confidence intervals (CIs) were estimated by the non-parametric bootstrap. Data analyses were performed using SPSS version 20.0 (IBM Corp., Armonk, NY).

## Results

### Demographic and clinical information

In the present study, in all participants, there were no significant differences in age between the ALS patients, LMNDCs and CIHCs, and age were significantly greater in AD patients and DDCs in our cohort. There were no significant differences in sex between the ALS patients, AD patients, LMNDCs, DDCs and CIHCs in our cohort. The demographic and clinical information of the whole cohort in our study is shown in Table 1, and the demographic and clinical information for each cohort is shown in Supplemental eTable 1-3.

**Table 1.** Demographic and Clinical Characteristics of the Pooled Cohort.

| Characteristic | AD patients<br>(n = 325) | ALS patients<br>(n = 235) | DDCs<br>(n = 145) | LMNDCs<br>(n = 289) | CIHCs<br>(n = 204) | *P* Value |
| --- | --- | --- | --- | --- | --- | --- |
| Age (years) | 68.46 ± 11.89 | 58.63 ± 11.52 | 67.59 ± 11.98 | 58.60 ± 11.31 | 58.25 ± 10.44 | <0.01 |
| Male/Female (n) | 195/130 | 139/96 | 87/58 | 174/115 | 123/81 | 0.99 |
| Disease duration (month) | - | 14.27 ± 10.23 | - | 14.12 ± 9.96 | - | 0.865 |
| ALSFRS-R score | - | 39.25 ± 7.06 | - | - | - | - |
| MMSE score | 22.13 ± 4.75 | 28.69 ± 1.76 | 23.50 ± 2.62 | 29.07 ± 0.87 | 29.22 ± 0.70 | <0.01 |
| MoCA score | 20.59 ± 4.26 | - | 20.90 ± 4.82 | - | 27.94 ± 1.40 | <0.01 |
| ECAS score | - | 93.56 ± 20.41 | - | - | 119.26 ± 8.95 | <0.01 |

Abbreviations: ALS = amyotrophic lateral sclerosis; CIHCs = cognitive intact healthy controls; ALSFRS-R = ALS Functional Rating Scale-Revised

### Serum brain-derived p-tau 217 tau levels alterations and their diagnostic performance in AD patients, ALS patients, DDCs, LMNDCs, and CIHCs

In the present study, in the whole cohort and each cohort, for serum brain-derived p-tau 217 levels, we found that serum brain-derived p-tau 217 levels were significantly higher in AD patients than ALS patients, DDCs, LMNDCs and, CIHCs after FWE correction (p < 0.05). There were no significant differences in serum brain-derived p-tau 217 levels among ALS patients, DDCs, LMNDCs and CIHCs. The profiles of serum brain-derived p-tau 217 alterations in all participants and each cohort are presented in Figure 2A-D.

**Figure 2.**
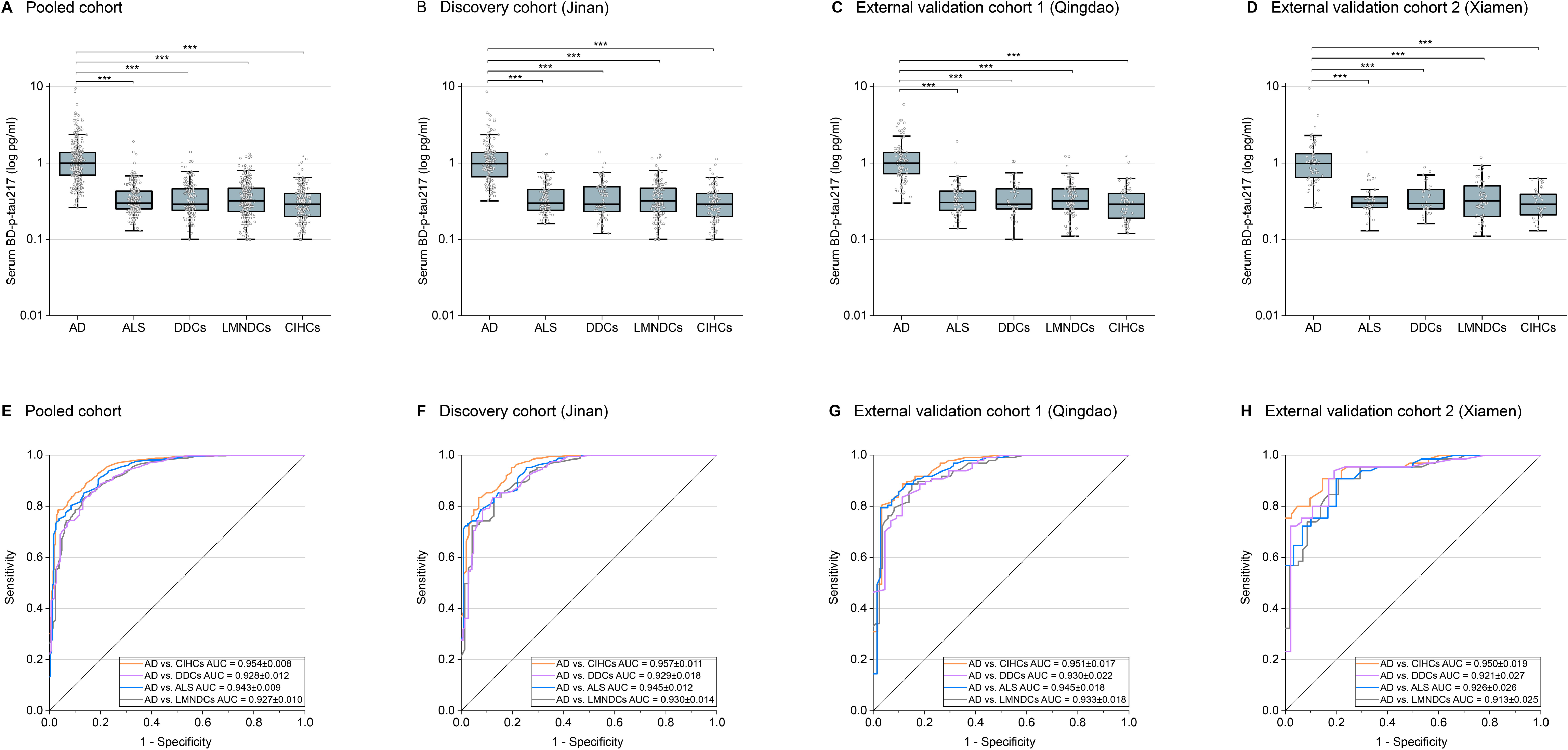
Serum Brain-Derived Phosphorylated Tau 217 Concentrations and Diagnostic Performance for Alzheimer Disease Across Cohorts. **A-D**, Box plots show serum BD–p-tau217 concentrations among participants with AD, ALS, DDCs, LMNDCs, and CIHCs in the pooled cohort (A), discovery cohort (Jinan) (B), external validation cohort 1 (Qingdao) (C), and external validation cohort 2 (Xiamen) (D). The horizontal line within each box indicates the median; the lower and upper bounds indicate the 25th and 75th percentiles, respectively; whiskers extend to the most extreme values within 1.5 times the interquartile range; and dots indicate individual participants. Concentrations are displayed on a logarithmic scale. Group differences were assessed using 1-way analysis of variance followed by pairwise comparisons with familywise error correction. Horizontal brackets indicate significant pairwise comparisons. \*\*\**P* < .001. **E-H,** ROC curves show the diagnostic performance of serum BD–p-tau217 in discriminating participants with AD from CIHCs, DDCs, participants with ALS, and LMNDCs in the pooled cohort (E), discovery cohort (F), external validation cohort 1 (G), and external validation cohort 2 (H). AUC estimates and corresponding SEs are shown in each panel. The diagonal reference line indicates discrimination no better than chance, corresponding to an AUC of 0.50. **Abbreviations:** AD, Alzheimer disease; ALS, amyotrophic lateral sclerosis; AUC, area under the receiver operating characteristic curve; BD–p-tau217, brain-derived phosphorylated tau 217; CIHC, cognitively intact healthy control; DDC, dementia control; LMNDC, lower motor neuron disease control; ROC, receiver operating characteristic; SE, standard error.

Moreover, in the whole samples, serum brain-derived p-tau 217 showed well diagnostic performance in discriminating AD patients from ALS patients (AUC 0.943±0.009), DDCs (AUC 0.928±0.012), LMNDCs (AUC 0.927±0.010), and CIHCs (AUC 0.954±0.008) (Figure 2E). Moreover, in Jinan cohort, serum brain-derived p-tau 217 also showed well diagnostic performance in discriminating AD patients from ALS patients (AUC 0.945±0.012), DDCs (AUC 0.929±0.018), LMNDCs (AUC 0.930±0.014), and CIHCs (AUC 0.957±0.011) (Figure 2F). In Qingdao cohort, serum brain-derived p-tau 217 also showed well diagnostic performance in discriminating AD patients from ALS patients (AUC 0.945±0.018), DDCs (AUC 0.930±0.022), LMNDCs (AUC 0.933±0.018), and CIHCs (AUC 0.951±0.017) (Figure 2G). In Xiamen cohort, serum brain-derived p-tau 217 also showed well diagnostic performance in discriminating AD patients from ALS patients (AUC 0.926±0.026), LMNDCs (AUC 0.913±0.025), and CIHCs (AUC 0.950±0.019) (Figure 2H).

### Serum total p-tau 217 levels alterations and their diagnostic performance in AD patients, ALS patients, DDCs, LMNDCs, and CIHCs

In the present study, in the whole samples and each cohort, for serum total p-tau 217 levels, we found that compared with CIHCs, DDCs and LMNDCs, serum total p-tau 217 levels were significantly higher in AD patients and ALS patients after FWE correction (p < 0.05). There were no significant differences in serum total p-tau 217 levels between LMNDCs, DDCs and CIHCs. The profiles of serum total p-tau 217 alterations in all participants are presented in Figure 3A-D.

**Figure 3.**
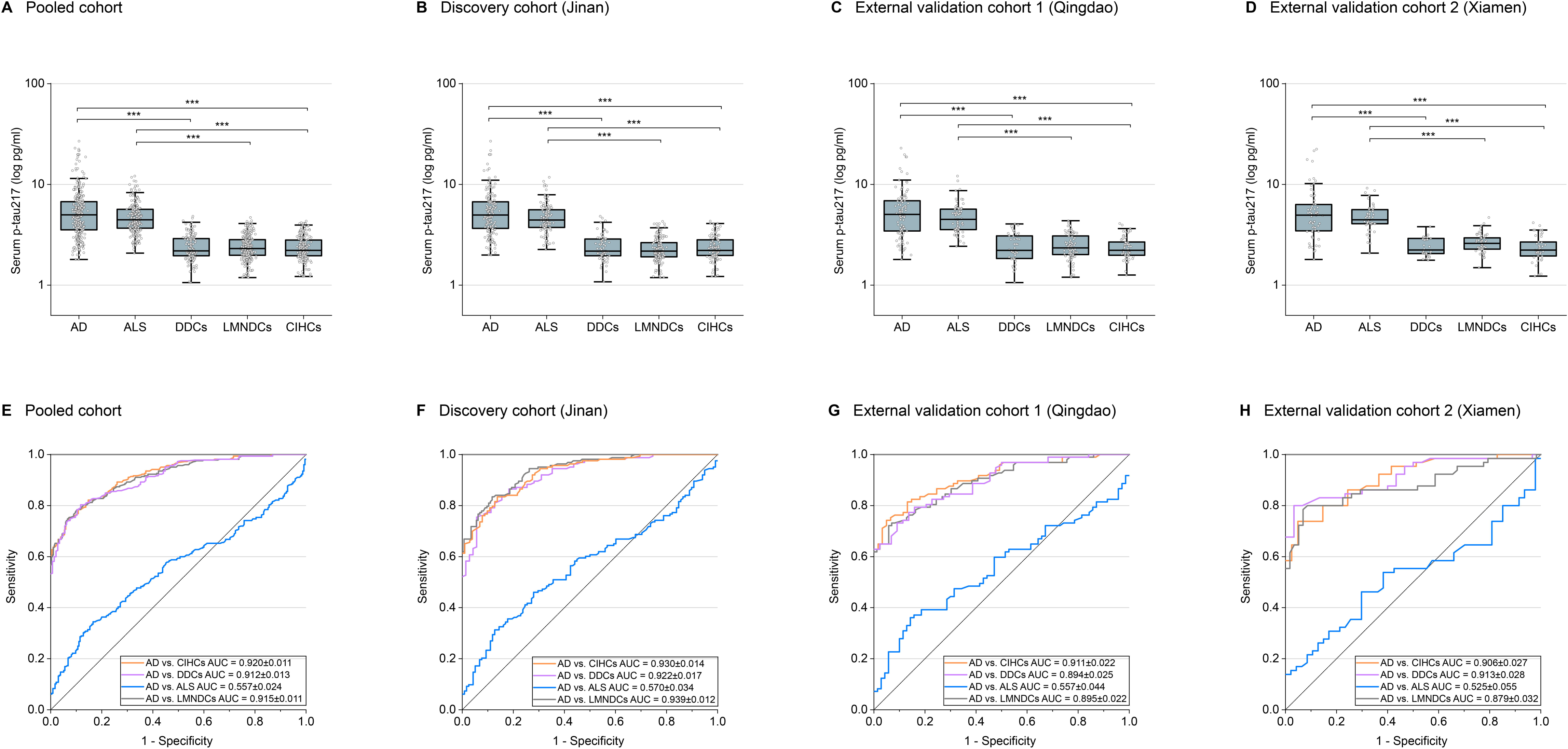
Serum Total p-tau217 Concentrations and Diagnostic Performance for AD Across Cohorts. **A-D**, Box plots show serum total p-tau217 concentrations among participants with AD, ALS, DDCs, LMNDCs, and CIHCs in the pooled cohort (A), discovery cohort (Jinan) (B), external validation cohort 1 (Qingdao) (C), and external validation cohort 2 (Xiamen) (D). The horizontal line within each box indicates the median; the lower and upper bounds indicate the 25th and 75th percentiles, respectively; whiskers extend to the most extreme values within 1.5 times the interquartile range; and dots indicate individual participants. Concentrations are displayed on a logarithmic scale. Group differences were assessed using 1-way analysis of variance followed by pairwise comparisons with familywise error correction. Horizontal brackets indicate significant pairwise comparisons; only significant comparisons are shown. \*\*\**P* < .001. **E-H,** ROC curves show the diagnostic performance of serum total p-tau217 in discriminating participants with AD from CIHCs, DDCs, participants with ALS, and LMNDCs in the pooled cohort (E), discovery cohort (F), external validation cohort 1 (G), and external validation cohort 2 (H). AUC estimates and corresponding SEs are shown in each panel. The diagonal reference line indicates discrimination no better than chance, corresponding to an AUC of 0.50. **Abbreviations:** AD, Alzheimer disease; ALS, amyotrophic lateral sclerosis; AUC, area under the receiver operating characteristic curve; CIHC, cognitively intact healthy control; DDC, dementia control; LMNDC, lower motor neuron disease control; p-tau217, phosphorylated tau at threonine 217; ROC, receiver operating characteristic; SE, standard error.

Moreover, in the whole samples, the diagnostic performance of serum total p-tau 217 in discriminating AD patients from ALS patients (AUC 0.557±0.024), LMNDCs (AUC 0.915±0.011), DDCs (AUC 0.912±0.013), and CIHCs (AUC 0.920±0.011) (Figure 3E). Moreover, in Jinan cohort, the diagnostic performance of serum total p-tau 217 in discriminating AD patients from ALS patients (AUC 0.570±0.034), LMNDCs (AUC 0.939±0.012), and CIHCs (AUC 0.930±0.014) (Figure 3F). In Qingdao cohort, the diagnostic performance of serum total p-tau 217 in discriminating AD patients from ALS patients (AUC 0.557±0.044), LMNDCs (AUC 0.895±0.022), and CIHCs (AUC 0.911±0.022) (Figure 3G). In Xiamen cohort, the diagnostic performance of serum total p-tau 217in discriminating AD patients from ALS patients (AUC 0.525±0.055), LMNDCs (AUC 0.879±0.032), and CIHCs (AUC 0.906±0.027) (Figure 3H).

To further compare the 2 p-tau217 measures, we examined the joint distribution of their log-transformed standardized values across the study groups. Patients with AD were characterized by elevations in both serum total p-tau217 and brain-derived p-tau217, whereas patients with ALS showed predominantly elevated total p-tau217 with comparatively lower brain-derived p-tau217 levels (eFigure 1 in the Supplement).

### Serum SV2A levels alterations and their diagnostic performance in AD patients, ALS patients, DDCs, LMNDCs, and CIHCs

In the present study, in all participants and each cohort, for serum SV2A levels, we found that compared with ALS patients, DDCs, CIHCs and LMNDCs, serum SV2A levels were significantly lower in AD patients after FWE correction (p < 0.05). The profiles of serum SV2A alterations in all participants and each cohort are presented in Figure 4A-D.

**Figure 4.**
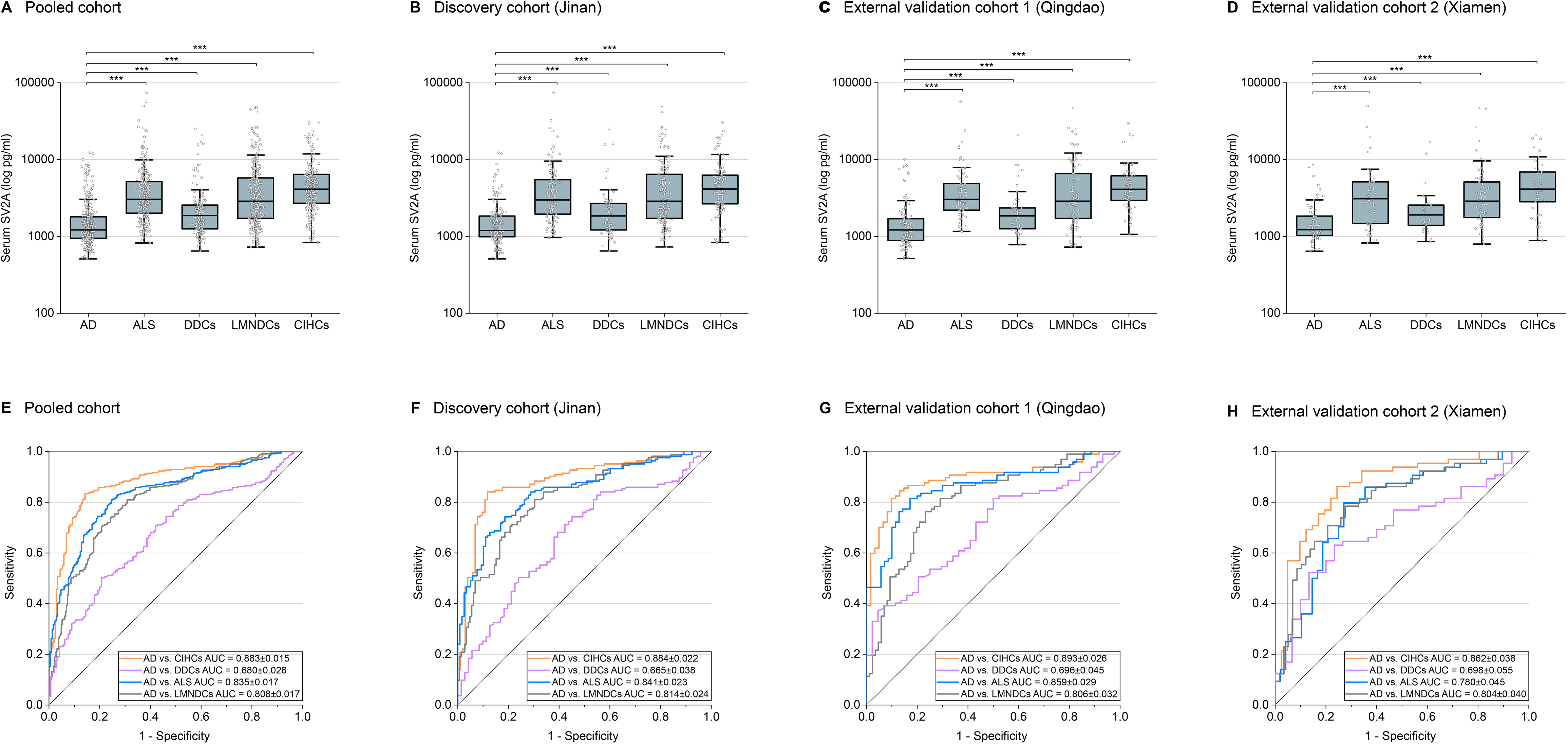
Serum SV2A Concentrations and Diagnostic Performance for AD Across Cohorts. **A-D**, Box plots show serum SV2A concentrations among participants with AD, ALS, DDCs, LMNDCs, and CIHCs in the pooled cohort (A), discovery cohort (Jinan) (B), external validation cohort 1 (Qingdao) (C), and external validation cohort 2 (Xiamen) (D). The horizontal line within each box indicates the median; the lower and upper bounds indicate the 25th and 75th percentiles, respectively; whiskers extend to the most extreme values within 1.5 times the interquartile range; and dots indicate individual participants. Concentrations are displayed on a logarithmic scale. Group differences were assessed using 1-way analysis of variance followed by pairwise comparisons with familywise error correction. Horizontal brackets indicate significant pairwise comparisons; only significant comparisons are shown. \*\*\**P* < .001. **E-H,** ROC curves show the diagnostic performance of serum SV2A in discriminating participants with AD from CIHCs, DDCs, participants with ALS, and LMNDCs in the pooled cohort (E), discovery cohort (F), external validation cohort 1 (G), and external validation cohort 2 (H). AUC estimates and corresponding SEs are shown in each panel. The diagonal reference line indicates discrimination no better than chance, corresponding to an AUC of 0.50. **Abbreviations:** AD, Alzheimer disease; ALS, amyotrophic lateral sclerosis; AUC, area under the receiver operating characteristic curve; CIHC, cognitively intact healthy control; DDC, dementia control; LMNDC, lower motor neuron disease control; ROC, receiver operating characteristic; SE, standard error; SV2A, synaptic vesicle glycoprotein 2A.

Finally, the diagnostic performance of serum SV2A in discriminating AD patients from ALS patients, LMNDCs, DDCs, and CIHCs also shown in Figure 4E-H. Receiver operating characteristic curve analysis was used to evaluate the diagnostic performance of the combination of serum brain-derived p-tau217 and SV2A for distinguishing patients with AD from those in the ALS, LMNDC, DDC, and CIHC groups (eFigure 2 in the Supplement).

### Serum NfL levels alterations and their diagnostic performance in AD patients, ALS patients, DDCs, LMNDCs, and CIHCs

In the present study, in all participants, for serum NfL levels, we found that compared with CIHCs, serum NfL levels were significantly higher in AD patients, ALS patients and LMNDCs after FWE correction (p < 0.05). Moreover, compared with AD and LMNDCs, serum NfL levels were significantly higher in ALS patients after FWE correction (p < 0.05). The profiles of serum NfL alterations among all participants are presented in eFigure 3A-D in the Supplement. The diagnostic performance of serum NfL is presented in eFigure 3E-H in the Supplement.

## Discussion

In a large multicentre cohort, we first found that blood brain-derived p-tau 217 and SV2A levels were promising brain damage biomarkers that can reduce peripheral damage interference and may be complementary for assisting early diagnosis in AD patients. Moreover, in this large multicentre cohort, we further suggest that blood total p-tau 217 is a specific biomarker for reflecting LMN damage in ALS patients. Thus, our findings may play crucial roles in advancing in early diagnosis and understanding the roles of p-tau species in AD patients and ALS patients.

In the present study, we first suggest that blood brain-derived p-tau 217 is likely a more specific biomarker for reflecting brain damage and may have less peripheral interference than blood total p-tau 217 in assisting in early diagnosis in AD patients.^7–18^ In our large multicentre study, we found that serum brain-derived p-tau 217 is significantly greater in AD patients than in ALS patients, LMNDCs, DDCs, and CIHCs, while serum p-tau 217 is significantly greater in both AD patients and ALS patients than in LMNDCs, DDCs, and CIHCs. Moreover, the AUC of serum brain-derived p-tau 217 for differentiating AD patients from ALS patients, DDCs, and CIHCs were better than the AUC of serum p-tau 217. To date, to our knowledge, very few studies have focused on serum brain-derived p-tau 217 alterations in AD patients.^16–18^ Recently, in a single centre head-to-head compared study, Jiang et al. aimed to compare the diagnostic performance of brain-derived p-tau 217 and total p-tau217 in 213 AD patients and 216 controls, and they found that brain-derived p-tau 217 have superior classification performance for AD pathology.^16^ Thus, they suggested that brain-derived p-tau 217 is likely a highly sensitive and specific blood-based biomarker for AD that has significant potential for early detection of AD-related brain damage.^16^ However, patients with peripheral damage were not included in that study.^16^ Thus, their results are strongly support and complementary with our study, these collect findings further underscore brain-derived p-tau 217 is likely a more ideal biomarker for AD patients in clinical practice and community screening.^16–18^ However, as mentioned above, at present, very few studies have focused on blood brain-derived p-tau 217 in AD patients; thus, further population and real world studies still need to confirm our findings.^16–18^

Moreover, our findings suggest that blood SV2A is a reliable biomarker and may reflect brain damage in AD patients. To date, to our knowledge, no large multicentre study has examined whether blood SV2A were altered and its diagnostic value in AD.^20^ Recently, in a single centre study, Wang et al. included 91 aMCI, 164 AD, 43 VaD (n = 43), 30 PDD, and 102 CIHCs, and blood SV2A, NfL, and p-tau 217 levels were measured.^20^ In that study, they found that blood SV2A demonstrated excellent diagnostic performance for aMCI, with a sensitivity of 97.8%, which was significantly higher than those of NfL and p-tau 217.^20^ Thus, they suggested that blood SV2A is likely a novel and ideal biomarker for the early diagnosis of AD.^20^ However, in that study, patients with peripheral damage were not included.^20^ Thus, our large multicentre findings may be complementary with their study and further suggest that blood SV2A is likely a promising brain damage biomarker for assessing synaptic damage in AD patients. Notably, in this study, we aimed to identify potential biomarker for reducing peripheral interference; thus, in addition to ALS, participants in the DDCs group were heterogeneous, further studies should be conducted to verify SV2A alterations and their roles in other specific neurodegenerative diseases, such as PD dementia and FTD in the future.^2–6^

Specifically, another key finding of our large multicentre study is that serum p-tau 217 is a potential diagnostic biomarker which can effectively differentiate ALS patients from other LMNDCs and CIHCs.^13–15^ Moreover, we found that serum p-tau 217 was not altered in LMNDCs in our cohorts. Thus, our study further suggests that serum p-tau 217 is a likely specific LMN damage biomarker and originate from muscle damage in ALS patient.^13^ To date, effectively biomarkers in assisting in early diagnosis in ALS patients remain largely lacking, and similar to 150 years ago when Jean-Martin Charcot first diagnostic this devastating disease, clinical evaluations remain the main approach for diagnosing ALS.^22^ Thus, the diagnosis delay for ALS patients can be as long as 8-15 months, and due to the heterogeneous of clinical symptoms in ALS patients, differentiating ALS from other LMNDCs can be difficult, in particular in early-stage ALS patients.^22, 34–37^

To date, only one large multicentre study has explored blood p-tau 217 abnormalities in ALS patients.^12–15^ Recently, in a multicentre study, Abu-Rumeileh et al. included 152 ALS patients, 111 AD patients, and 99 non-neurodegenerative disease controls, and they found that serum p-tau 217 levels were increased in both AD patients and ALS patients than in controls.^12^ In that study, they found that the elevation of blood p-tau 217 in ALS patients is likely to reflect the release of p-tau 217 from denervated muscle fibres.^12^ Specifically, in that study, other LMN damage diseases were not included, and the authors also pointed to the need for further studies to explore serum p-tau 217 in other neuromuscular diseases.^12^ In our cohorts, we found that, compared with CIHCs, serum p-tau 217 levels were significantly increased in ALS patients and were not altered in LMNDCs. Thus, our study continues to support their findings and further suggest that serum p-tau 217 is likely a specific LMN damage biomarker for ALS patients and can effectively differentiate ALS patients from other neuromuscular diseases.^12^ Importantly, although EMG can effectively detect LMN damage in ALS, EMG is time-consuming, and some patients find it difficult to tolerate the pain. Thus, serum p-tau 217 may provide the benefits of increased convenience and non-invasiveness for evaluating LMN damage in ALS.^12–15^ However, as mentioned above, at present, very few studies have focused on blood p-tau 217 alterations in ALS patients; thus, further studies still need to confirm our viewpoints.^12–15^

Overall, in a large multicentre cohort, we demonstrated that blood brain-derived p-tau 217 was not altered in patients with ALS or other LMN diseases and we suggest that brain-derive p-tau 217 and SV2A are likely more ideal biomarkers that can reduce peripheral interference in assisting early diagnosis for AD patients.^6–15^ Specifically our findings further suggest that serum p-tau 217 is a specific LMN damage biomarker and can originate from muscle damage in ALS patients. Thus, our findings may play crucial roles in advancing in early diagnosis for AD patients and ALS patients. However, inevitably, our study had several limitations. First, although the total sizes of our cohort were relatively large, further population study is still needed to verify our findings. However, the clinical and epidemiological data of the AD patients and ALS patients were similar to those of recent national population-based studies, which may thus strengthen the validity and credibility of our findings.^38–40^ Second, we only use SMID approach to assess brain-derived p-tau 217 and SV2A levels.^16–18^ Further studies are still needed to use other approach, such as NULISA, to verify serum brain-derived p-tau 217 and SV2A levels in the future. Finally, in this study, we only included Chinese participants, further studies are still needed to verify our findings in other races AD patients and ALS patients in the future.

In conclusion, our findings suggest that blood brain-derived p-tau 217 is a more ideal biomarker for reflecting brain damage and may not be disturbed by peripheral damage; thus, blood brain-derived p-tau 217 combined with SV2A may provide more effective diagnostic performance for AD patients. Moreover, we suggest that serum p-tau 217 is a specific LMN damage biomarker for ALS patients. However, further studies are still needed to confirm our findings.

## Supporting information

Supplementary STROBE Checklist

## Acknowledgements

We thank all participants.

## Funding

This work was supported by the Key R&D Program of Shandong Province, China (Grant No: 2025CXPT133) and the National natural science foundation of China (Grant number: No. 82471429, 82071412 and 82171395).

## Conflicts of Interest

No.

## Data Availability

The anonymized data presented in this article are available at the request of a qualified investigator, after review by the corresponding authors. Final approval will be granted by the Research Ethics Committee of the Qilu Hospital, Shandong University.

## Author Contributions

SWL: study concepts and write the first version of the manuscript. MJM, HXW, XHL JYL and NLG: interpretation of the results, revising the manuscript. FCL: study concepts, interpretation of the results. JL, XHS, KLW, OC, JWX and KS: revising the manuscript. WL and YYZ: data acquisition, revising the manuscript. CZY, YL, and FCL: diseases diagnosis and revising the manuscript.

## Notes

### Competing Interest Statement

The authors have declared no competing interest.

### Author Declarations

The Institutional Review Board of Qilu Hospital of Shandong University gave ethical approval for this work (approval number: KYLL-202412-067-1).

## References

1. Mielke MM, Fowler NR. Alzheimer disease blood biomarkers: considerations for population-level use. Nat Rev Neurol. 2024 Aug;20(8):495–504.

2. Hansson O, Blennow K, Zetterberg H, Dage J. Blood biomarkers for Alzheimer’s disease in clinical practice and trials. Nat Aging. 2023 May;3(5):506–519.

3. Jack CR Jr, Andrews JS, Beach TG, et al. Revised criteria for diagnosis and staging of Alzheimer’s disease: Alzheimer’s Association Workgroup. Alzheimers Dement. 2024 Aug;20(8):5143–5169.

4. Howe MD, Britton KJ, Joyce HE, et al. Clinical application of plasma P-tau217 to assess eligibility for amyloid-lowering immunotherapy in memory clinic patients with early Alzheimer’s disease. Alzheimers Res Ther. 2024 Jul 6;16(1):154.

5. Teunissen CE, Kolster R, Triana-Baltzer G, Janelidze S, Zetterberg H, Kolb HC. Plasma p-tau immunoassays in clinical research for Alzheimer’s disease. Alzheimers Dement. 2025 Jan;21(1):e14397.

6. Mielke MM, Anderson M, Ashford JW, et al. Recommendations for clinical implementation of blood-based biomarkers for Alzheimer’s disease. Alzheimers Dement. 2024 Nov;20(11):8216–8224.

7. Ashton NJ, Brum WS, Di Molfetta G, et al. Diagnostic Accuracy of a Plasma Phosphorylated Tau 217 Immunoassay for Alzheimer Disease Pathology. JAMA Neurol. 2024 Mar 1;81(3):255–263.

8. Gao F, Dai L, Wang Q, et al. Blood-based biomarkers for Alzheimer’s disease: a multicenter-based cross-sectional and longitudinal study in China. Sci Bull (Beijing). 2023 Aug 30;68(16):1800–1808.

9. Jiao B, Ouyang Z, Liu Y, et al. Evaluating the diagnostic performance of six plasma biomarkers for Alzheimer’s disease and other neurodegenerative dementias in a large Chinese cohort. Alzheimers Res Ther. 2025 Apr 3;17(1):71.

10. Mielke MM, Dage JL, Frank RD, et al. Performance of plasma phosphorylated tau 181 and 217 in the community. Nat Med. 2022 Jul;28(7):1398–1405.

11. Fernández Arias J, Brum WS, Salvadó G, et al. Plasma phosphorylated tau217 strongly associates with memory deficits in the Alzheimer’s disease spectrum. Brain. 2025 Jul 7;148(7):2384–2399.

12. Abu-Rumeileh S, Scholle L, Mensch A, et al. Phosphorylated tau 181 and 217 are elevated in serum and muscle of patients with amyotrophic lateral sclerosis. Nat Commun. 2025 Mar 5;16(1):2019.

13. Thomas EV, Han C, Kim WJ, et al. ALS plasma biomarkers reveal neurofilament and pTau correlate with disease onset and progression. Ann Clin Transl Neurol. 2025 Apr;12(4):714–723.

14. Vacchiano V, Mastrangelo A, Zenesini C, et al. Elevated plasma p-tau181 levels unrelated to Alzheimer’s disease pathology in amyotrophic lateral sclerosis. J Neurol Neurosurg Psychiatry. 2023 Jun;94(6):428–435.

15. Bozkurt H, Reid KR, Newton J, et al. Blood-based biomarker discovery in motor neuron disease using nucleic acid-linked immuno-sandwich assay. Brain Commun. 2026 May 26;8(3):fcag180.

16. Jiang Y, Zheng W, Xia Z, et al. Head-to-head comparison of brain-derived pTau217 and total pTau217 for brain amyloid and tau pathology classification. Proc Natl Acad Sci U S A. 2026 Mar 10;123(10):e2536792123.

17. Ghisays V, Denkinger MN, Singh A, et al. Prognostic value of plasma brain-derived pTau. medRxiv [Preprint]. 2026 Jun 30:2026.06.26.26356597.

18. Chong JR, Hilal S, Venketasubramanian N, et al. Plasma brain-derived p-Tau217 outperforms other p-Tau species in detecting abnormal brain amyloid in an Asian cohort of older people with cerebrovascular disease burden. J Prev Alzheimers Dis. 2026 Aug;13(7):100615.

19. Jack CR Jr, Bennett DA, Blennow K, et al. NIA-AA Research Framework: Toward a biological definition of Alzheimer’s disease. Alzheimers Dement. 2018 Apr;14(4):535–562.

20. Wang X, Zhang X, Liu J, et al. Synaptic vesicle glycoprotein 2 A in serum is an ideal biomarker for early diagnosis of Alzheimer’s disease. Alzheimers Res Ther. 2024 Apr 13;16(1):82.

21. Mecca AP, Ashton NJ, Chen MK, et al. Cerebrospinal fluid and brain positron emission tomography measures of synaptic vesicle glycoprotein 2A: Biomarkers of synaptic density in Alzheimer’s disease. Alzheimers Dement. 2025 Jun;21(6):e70344.

22. Irwin KE, Sheth U, Wong PC, Gendron TF. Fluid biomarkers for amyotrophic lateral sclerosis: a review. Mol Neurodegener. 2024 Jan 24;19(1):9.

23. Lin J, Liu Y, Ouyang Z, et al. Multidimensional modeling of biological aging: integrating gait, eye movement, rest-state functional connectivity, and plasma biomarkers in non-dementia older adults. J Prev Alzheimers Dis. 2026 Jun;13(6):100566.

24. Xiao J, Li B, Li B, et al. A prospective real-world study of the efficacy and safety of aducanumab in China: Focus on early-onset and autosomal dominant Alzheimer’s disease. Alzheimers Dement (Amst). 2026 Apr 8;18(2):e70328.

25. Jiao B, Ouyang Z, Xiao X, et al. Development and validation of machine learning models with blood-based digital biomarkers for Alzheimer’s disease diagnosis: a multicohort diagnostic study. EClinicalMedicine. 2025 Mar 5;81:103142.

26. de Carvalho M, Dengler R, Eisen A, et al. Electrodiagnostic criteria for diagnosis of ALS. Clin Neurophysiol. 2008 Mar;119(3):497–503.

27. Liu S, Sun X, Ren Q, et al. Glymphatic dysfunction in patients with early-stage amyotrophic lateral sclerosis. Brain. 2024 Jan 4;147(1):100–108.

28. Chen Y, Sun S, Bai Z, et al. CSF Aβ, Tau, Axonal, Synaptic, Glial, Neural, and Inflammatory Biomarkers in Patients With Sporadic Amyotrophic Lateral Sclerosis. Neurology. 2025 Oct 7;105(7):e213914.

29. McKhann GM, Knopman DS, Chertkow H, et al. The diagnosis of dementia due to Alzheimer’s disease: recommendations from the National Institute on Aging-Alzheimer’s Association workgroups on diagnostic guidelines for Alzheimer’s disease. Alzheimers Dement. 2011 May;7(3):263–9.

30. Thompson AG, Gray E, Bampton A, Raciborska D, Talbot K, Turner MR. CSF chitinase proteins in amyotrophic lateral sclerosis. J Neurol Neurosurg Psychiatry. 2019 Nov;90(11):1215–1220.

31. Kwan J, Vullaganti M. Amyotrophic lateral sclerosis mimics. Muscle Nerve. 2022 Sep;66(3):240–252.

32. Obara K, Ito D, Nilsson C, Janelidze S, Santillo A, Katsuno M, Mattsson-Carlgren N. Diagnostic and Prognostic Value of Blood and Cerebrospinal Fluid Biomarkers in Amyotrophic Lateral Sclerosis: A Systematic Review and Meta-Analysis. Eur J Neurol. 2025 Oct;32(10):e70382.

33. Chen Y, Sun S, Gao N, et al. Proximity extension assay reveals serum inflammatory biomarkers in two amyotrophic lateral sclerosis cohorts. Neurobiol Dis. 2025 Jul;211:106933.

34. Gwathmey KG, Corcia P, McDermott CJ, Genge A, Sennfält S, de Carvalho M, Ingre C. Diagnostic delay in amyotrophic lateral sclerosis. Eur J Neurol. 2023 Sep;30(9):2595–2601.

35. Taylor JP, Brown RH Jr, Cleveland DW. Decoding ALS: from genes to mechanism. Nature. 2016 Nov 10,539(7628):197–206.

36. Kiernan MC, Vucic S, Talbot K, et al. Improving clinical trial outcomes in amyotrophic lateral sclerosis. Nat Rev Neurol. 2021 Feb;17(2):104–118.

37. Al-Chalabi A, Calvo A, Chio A, et al. Analysis of amyotrophic lateral sclerosis as a multistep process: a population-based modelling study. Lancet Neurol. 2014 Nov;13(11):1108–1113.

38. Xu L, Chen L, Wang S, et al. Incidence and prevalence of amyotrophic lateral sclerosis in urban China: a national population-based study. J Neurol Neurosurg Psychiatry. 2020 May;91(5):520–525.

39. Tian X, Zhu M, Ma Y, et al. Physical and biopsychosocial frailty, cognitive phenotypes, and plasma biomarkers for Alzheimer’s disease in Chinese older adults: A population-based study. Alzheimers Dement. 2025 May;21(5):e70303.

40. Liu MS, Cui LY, Fan DS; Chinese ALS Association. Age at onset of amyotrophic lateral sclerosis in China. Acta Neurol Scand. 2014 Mar;129(3):163–7.

