## Supplementary figures and images for "Serum brain-derived p-tau 217 and SV2A Reduce Peripheral Interference in Alzheimer’s Disease: A Multicohort Study"

### Supplementary Figure 1

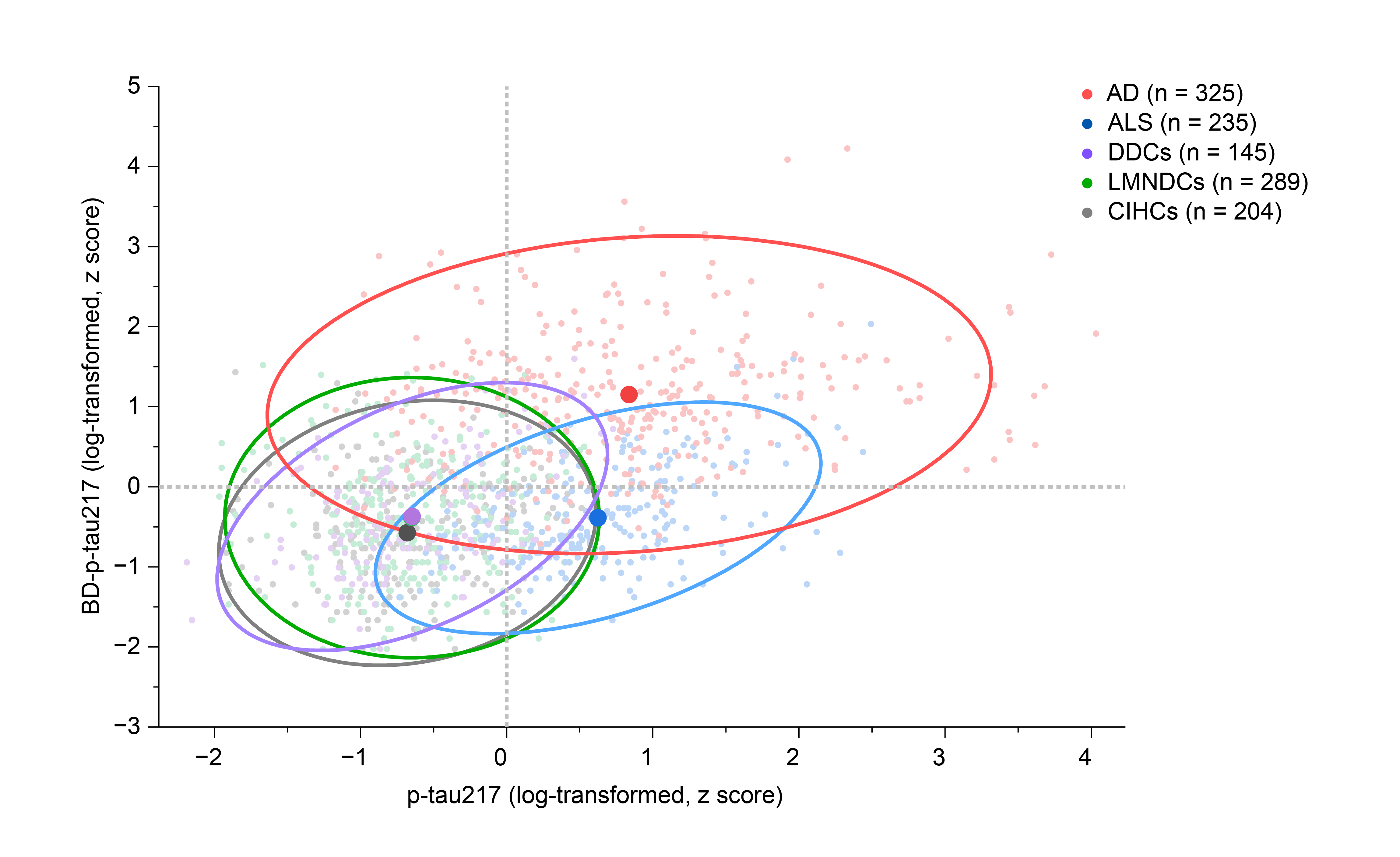

### Supplementary Figure 2

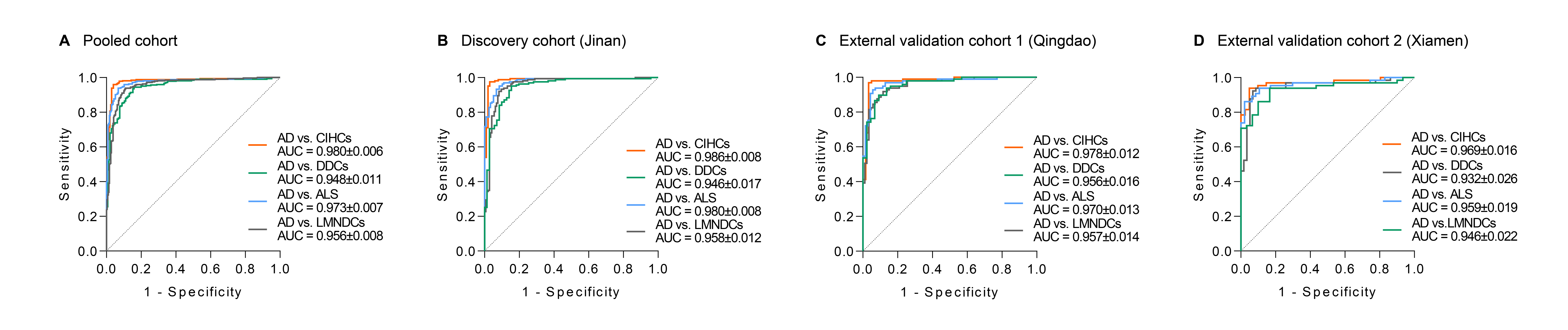

### Supplementary Figure 3

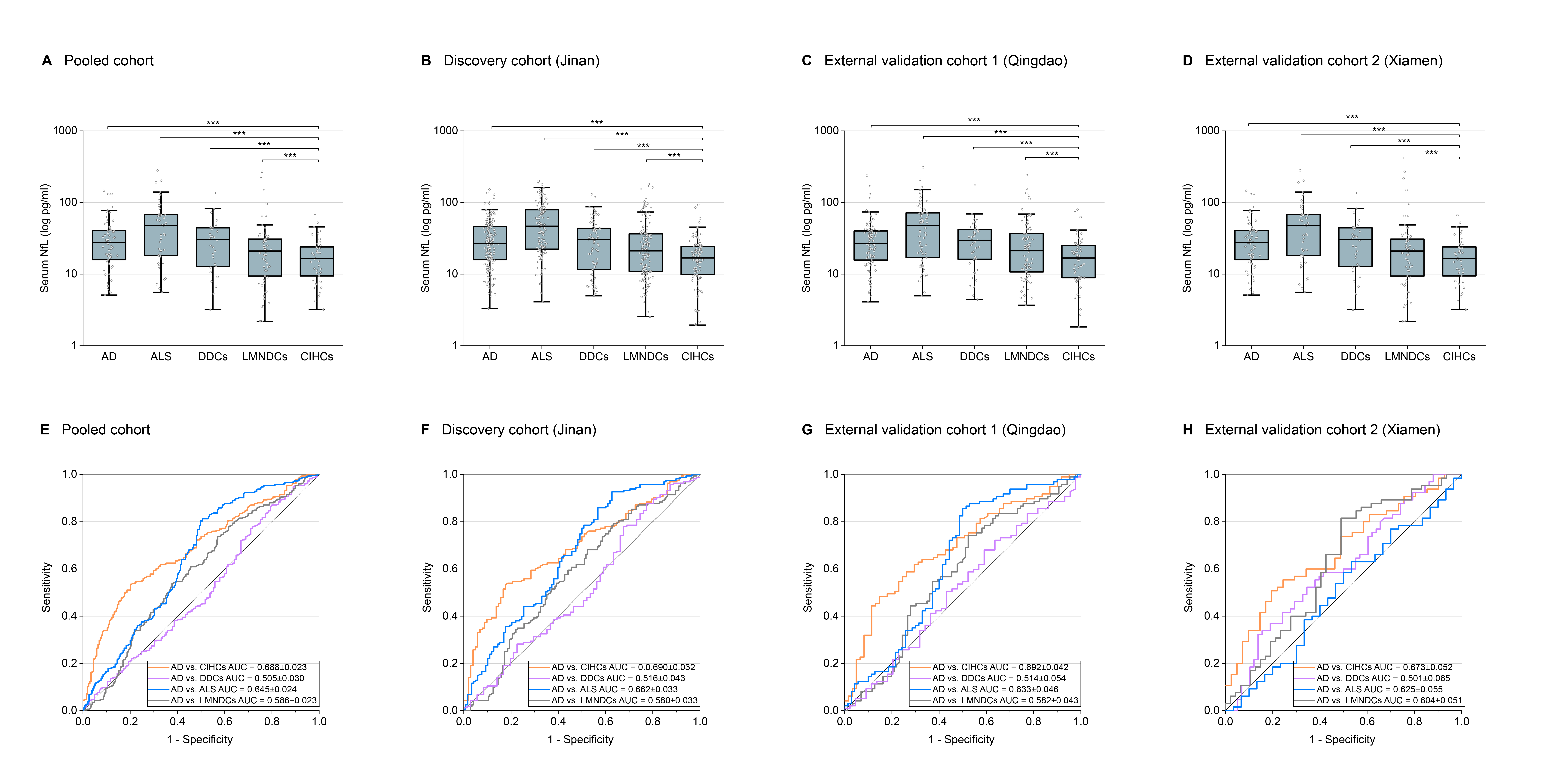
