## Supplementary material for "Serum brain-derived p-tau 217 and SV2A Reduce Peripheral Interference in Alzheimer’s Disease: A Multicohort Study"

eTable 1. Demographic and Clinical Characteristics of the Discovery Cohort (Jinan)

| **Characteristic** | AD patients (n = 163) | ALS patients (n = 118) | DDCs (n = 71) | LMNDCs (n = 145) | CIHCs (n = 102) | *P* Value |
| --- | --- | --- | --- | --- | --- | --- |
| Age (years) | 68.89 ± 12.72 | 58.39 ± 11.96 | 67.39 ± 10.86 | 57.77 ± 12.01 | 58.16 ± 10.84 | <0.01 |
| Male/Female (n) | 98/65 | 71/47 | 43/28 | 87/58 | 61/41 | 0.99 |
| Disease duration (month) | - | 14.92 ± 9.95 | - | 14.72 ± 10.03 | - | 0.873 |
| ALSFRS-R score | - | 38.90 ± 7.46 | - | - | - | - |
| MMSE score | 22.16 ± 4.67 | 28.49 ± 1.90 | 23.75 ± 2.51 | 29.12 ± 0.88 | 29.23 ± 0.76 | <0.01 |
| MoCA score | 19.99 ± 4.07 | - | 20.38 ± 4.81 | - | 28.03 ± 1.38 | <0.01 |
| ECAS score | - | 89.92 ± 21.48 | - | - | 119.48 ± 9.04 | <0.01 |

eTable 2. Demographic and Clinical Characteristics of the External Validation Cohort 1 (Qingdao)

| **Characteristic** | AD patients (n = 97) | ALS patients (n = 70) | DDCs (n = 44) | LMNDCs (n = 86) | CIHCs (n = 61) | *P* Value |
| --- | --- | --- | --- | --- | --- | --- |
| Age (years) | 67.87 ± 10.06 | 59.21 ± 10.12 | 67.23 ± 11.54 | 59.44 ± 9.87 | 58.21 ± 10.01 | <0.01 |
| Male/Female (n) | 58/39 | 40/30 | 26/18 | 52/34 | 37/24 | 0.994 |
| Disease duration (month) | - | 14.09 ± 11.43 | - | 13.80 ± 10.69 | - | 0.874 |
| ALSFRS-R score | - | 39.16 ± 7.91 | - | - | - | - |
| MMSE score | 21.63 ± 5.07 | 28.86 ± 1.69 | 22.91 ± 2.56 | 29.01 ± 0.87 | 29.10 ± 0.68 | <0.01 |
| MoCA score | 20.77 ± 4.30 | - | 21.45 ± 4.57 | - | 27.95 ± 1.37 | <0.01 |
| ECAS score | - | 95.74 ± 18.94 | - | - | 120.16 ± 9.47 | <0.01 |

eTable 3. Demographic and Clinical Characteristics of External Validation Cohort 2 (Xiamen)

| **Characteristic** | AD patients (n=65) | ALS patients (n=47) | DDCs (n = 30) | LMNDCs (n = 58) | CIHCs (n = 41) | *P* Value |
| --- | --- | --- | --- | --- | --- | --- |
| Age (years) | 68.28 ± 12.38 | 58.34 ± 12.52 | 68.60 ± 15.15 | 59.41 ± 11.52 | 58.51 ± 10.30 | <0.01 |
| Male/Female (n) | 39/26 | 28/19 | 18/12 | 35/23 | 25/16 | 0.99 |
| Disease duration (month) | - | 12.91 ± 8.99 | - | 13.09 ± 8.62 | - | 0.921 |
| ALSFRS-R score | - | 40.26 ± 4.09 | - | - | - | - |
| MMSE score | 22.80 ± 4.43 | 28.91 ± 1.49 | 23.80 ± 2.92 | 29.05 ± 0.85 | 29.39 ± 0.54 | <0.01 |
| MoCA score | 21.85 ± 4.42 | - | 21.30 ± 5.22 | - | 27.71 ± 1.50 | <0.01 |
| ECAS score | - | 99.47 ± 18.18 | - | - | 117.39 ± 7.80 | <0.01 |

**eFigure 1. Joint Distribution of Serum Total and Brain-Derived Phosphorylated Tau 217 Across Diagnostic Groups**

Each point represents an individual participant. Larger points indicate group means, and ellipses summarize the distribution of each group. Biomarker concentrations were log-transformed and standardized as z scores.

**Abbreviations:** AD, Alzheimer disease; ALS, amyotrophic lateral sclerosis; BD–p-tau217, brain-derived phosphorylated tau 217; CIHC, cognitively intact healthy control; DDC, dementia disease control; LMNDC, lower motor neuron disease control.

**eFigure 2. Diagnostic Performance of Combined Serum Brain-Derived Phosphorylated Tau 217 and SV2A**

Receiver operating characteristic curves show discrimination of AD from CIHCs, DDCs, ALS, and LMNDCs in the pooled cohort (A), discovery cohort (B), external validation cohort 1 (C), and external validation cohort 2 (D). AUC values and SEs are shown.

**Abbreviations:** AD, Alzheimer disease; ALS, amyotrophic lateral sclerosis; AUC, area under the curve; CIHC, cognitively intact healthy control; DDC, dementia disease control; LMNDC, lower motor neuron disease control; SV2A, synaptic vesicle glycoprotein 2A.

**eFigure 3. Serum Neurofilament Light Chain Concentrations and Diagnostic Performance Across Cohorts**

A-D, Serum NfL concentrations across diagnostic groups in the pooled, discovery, and 2 external validation cohorts. E-H, ROC curves for distinguishing AD from CIHCs, DDCs, ALS, and LMNDCs. AUC values and SEs are shown. ****P* < .001.

**Abbreviations:** AD, Alzheimer disease; ALS, amyotrophic lateral sclerosis; AUC, area under the curve; CIHC, cognitively intact healthy control; DDC, dementia disease control; LMNDC, lower motor neuron disease control; NfL, neurofilament light chain; ROC, receiver operating characteristic.
